# Preventing postpartum depression through mitigating breastfeeding grief: A convergent parallel mixed methods study

**DOI:** 10.64898/2026.06.08.26354940

**Authors:** Tumilara Aderibigbe, Clarisa Medina-Poeliniz, Ryoko Kausler, Blessing Okorie, Sara E. Simonsen, Gwen Latendresse

## Abstract

**Background:** Women who did not meet their breastfeeding goals often experience breastfeeding grief (BG) and may be likely to have postpartum depression (PD). Furthermore, PD is nearly twice as common in African American (AA) women as in Non-Hispanic White women. No research exists on BG and its role in PD. This study examined AA women’s experiences of BG and its possible contributions to PD symptoms.

**Methods:** A convergent parallel mixed methods design was used. A purposive sample of 16 AA women with children aged 6 months to 2 years with BG participated in individual semi-structured interviews about their experiences of BG and completed an online survey including the Edinburgh Postnatal Depression Scale (EPDS). Qualitative and quantitative data were analyzed using reflexive thematic analysis and descriptive statistics, respectively. Both data were integrated using joint display of data and side-by-side comparison.

**Results:** Participants’ mean age was 29.5 years. Four meaning-based themes about BG were generated including: *We looked forward to breastfeeding, But it did not go as expected*, *So we grieve*, and *These would have helped.* From quantitative results, 87.5% of participants reported a history of PD symptoms and almost 44% had EPDS scores >11. All participants reported that experiencing BG contributed to their PD symptoms. Findings suggest that BG influenced PD symptoms in AA women without prior diagnosis of depression.

**Conclusions:** Qualitative and quantitative findings from this novel exploratory study revealed an overlap that AA women with BG report PD symptoms. Clinicians should support women to achieve their breastfeeding goals to prevent BG and PD.

## Introduction

The 140% increase in the prevalence of postpartum depression (PD) among African American (AA) women in the United States from 2010 to 2021 is a public health concern (Getahun et al., 2023). Postpartum depression negatively impacts the mental health and overall functioning of AA women (Floyd James et al., 2023). In an attempt to uncover and prevent PD in AA women, some factors influencing PD in AA women were identified including low socioeconomic status (Dolbier et al., 2013), teenage or first-time mother (Ceballos et al., 2017), cesarean or episiotomy site pain (Howell et al., 2005), neighborhood disadvantage (Onyewuenyi et al., 2023), lack of social support (Cannon & Nasrallah, 2019), and diagnosis of prepartum depression (Ceballos et al., 2017; Liu & Tronick, 2013). Yet, AA women continue to be twice as likely to have PD symptoms compared to Non-Hispanic White women (Tabb et al., 2020). Therefore, further examination of other possible risk factors for PD is crucial to prevent PD in AA women and promote maternal mental well-being.

Breastfeeding grief (BG), experienced by women who did not meet their breastfeeding goals, is defined as profound (and often prolonged) sadness and a sense of failure that negatively impacts maternal psychological well-being (Author, 2026; Brown, 2019). Among children born in 2022, 78% of Non-Hispanic Black infants were ever breastfed. However, only 54%, 33%, and 9% were breastfed for 6 months, 12 months, and 2 years postpartum, respectively (Centers for Disease Control and Prevention, 2022). Although BG may contribute to PD, this relationship remains unexplored (Demirci, 2022). Previous qualitative studies have documented depressive symptoms among women experiencing BG (Brown, 2018; Gribble et al., 2023; Robinson, 2018). However, quantitative research is needed to understand the relationship between BG and PD. Our study addressed this research gap using a mixed methods approach (i.e. integration of qualitative and quantitative methods of data collection and analysis) (Creswell, 1999; Creswell & Creswell, 2018) to explore the possible contributions of BG to PD. The research questions in line with Tashakkori and Creswell (2007)’s guidelines (Tashakkori & Creswell, 2007) were:

1. How do African American women experience breastfeeding grief and what are the perceived barriers and resources needed to achieve their breastfeeding goals (qualitative)?
2. Do African American women with breastfeeding grief have past or current postpartum depressive symptoms (quantitative)?
3. Do quantitative results about breastfeeding grief and postpartum depressive symptoms converge with qualitative findings (mixed methods)?

## Materials and Methods

### Philosophical and Theoretical Framework

This study has its philosophical underpinning in Pragmatism (Cherryholmes, 1992). Applied to mixed methods research, pragmatism enables researchers to investigate a research problem using different methods of data collection and analysis (Creswell & Creswell, 2018; Morgan, 2007; Patton, 1990). Constructionist epistemology (Braun & Clarke, 2019; Byrne, 2022) guided the generation of meaning-based themes of BG.

### Study Design

Convergent parallel mixed methods design (Creswell & Plano Clark, 2011; Creswell & Creswell, 2018) was used to compare perspectives of AA women about BG and its contributions to PD, to examine if qualitative and quantitative findings confirm or disconfirm each other in this exploratory study (Figure 1; adopted from (Author et al., 2026)). Though qualitative and quantitative data were collected concurrently, the qualitative strand is the dominant strand of the study (i.e QUAL + quant (Morse, 1991)) because the concept of BG is “immature” (Morse, 1991) (p.120) or understudied, and we intended to validate qualitative findings about BG and PD symptoms using quantitative data. Descriptive design was used for the qualitative strand, as the most suitable qualitative approach for a mixed methods study (Neergaard et al., 2009), especially when little is known about the phenomenon examined (Doyle et al., 2020; Sandelowski, 2010). Descriptive cross-sectional design was used for the quantitative strand (Aggarwal & Ranganathan, 2019).

**Figure 1.**
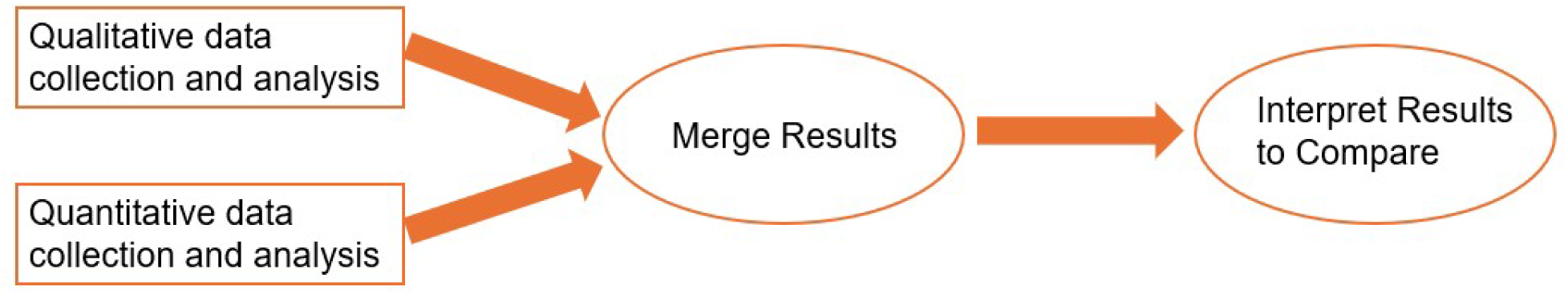
Convergent Parallel Mixed Methods Design. Adopted from (Aderibigbe et al., 2026)

### Sample

The qualitative strand is the dominant strand of the study hence we enrolled a purposive sample of 16 women. A minimum sample size of 9 participants is required to reach data saturation using interviews (Hennink & Kaiser, 2022). We included women who: (a) self-identify as AA woman; (b) are 18 years or older; (c) have access to the internet; (d) can speak and read English; (e) are birth mothers to a child aged 6 months-2 years with full-term gestation; (f) had a singleton pregnancy; (g) initiated breastfeeding; and (h) are experiencing/experienced BG (i.e. profound sadness due to unmet breastfeeding goal). Women with children up to 2 years were included because of WHO’s recommendation to breastfeed for at least 2 years (World Health Organization, 2026) and reports from earlier studies of prolonged BG up to 36 years postpartum (Brown, 2019) and prolonged PD between 3-11 years postpartum (Netsi et al., 2018; Putnick et al., 2020). Women who: (a) were < 18 years of age; (b) have a diagnosis of serious mental health disorder e.g., schizophrenia, bipolar disorder; and (c) have breast conditions that may impact breastfeeding (e.g. mastectomy) were excluded.

### Setting

Passive online recruitment using a virtual research flyer was conducted for 1 month. Virtual research flyers were posted on Facebook and LinkedIn to target a national sample of breastfeeding AA women in the United States.

### Ethical considerations

The study was approved by the University of XX Institutional Review Board (IRB#: 00176280). A waiver of informed consent documentation was granted because the study poses no more than minimal risk to participants,. An electronic informed consent was provided to individuals who wish to participate. TA reviewed informed consent with participants and answered their questions. Interested participants were thereafter enrolled in the study after informed consent was granted. To maintain confidentiality and data security, data were stored on the University of XX’s secure cloud storage and only the research team had access to the data (University of XXX, 2023).

### Data Collection

The research flyer contained a QR code to the link for a Research Electronic Data Capture (REDCap) screening survey and TA’s email address. Participants either contacted TA via email to indicate interest or completed screening and were followed up. Eligible participants identified from the screening survey were contacted via email to schedule the interview and TA obtained informed consent from interested participants.

### Qualitative

#### Reflexivity

Reflexivity refers to identifying researchers’ positioning, and how it shaped different sections of the study (Braun & Clarke, 2023; Trainor & Bundon, 2021). TA, CM, and BO have previously conducted research on breastfeeding and factors preventing women from meeting their breastfeeding goals among AA and Hispanic women (Author & Lucas, 2022; Author et al., 2023; Stanhope et al., 2024). RK, SS, and GL have also conducted research on PD (Latendresse et al., 2023). We consciously examined and put aside our premonitions and findings from previous studies throughout the study. Further, we engaged in introspection reflexivity, intersubjective reflection, and mutual collaboration (Finlay, 2002; Trainor & Bundon, 2021). Details about how reflexivity was maintained throughout the study (Finlay, 2002; Trainor & Bundon, 2021) are included in Supplementary material 1.

Utilizing the definition of BG from our previous article (Author, 2026), BG was defined in the current study as profound (and often prolonged) sadness and a sense of failure that negatively impacts maternal psychological well-being. Semi-structured individual interviews were conducted because BG is a sensitive topic, and we intended to have a personalized and private discussion while gathering in-depth data from individual participants without external influence (Kruger et al., 2019). Additional information about qualitative data collection were presented in our previous article (Author, 2026). ***Quantitative:*** Before the interview commenced, participants completed an online survey on REDCap which took approximately 10 minutes. The survey included items on demographics, lactation, and the Edinburgh Postnatal Depression Scale (EPDS), which contains 10 items on a 4-point Likert scale and has a reliability score of 0.88 (Cox et al., 1987). The EPDS is valid and reliable to measure PD symptoms in the perinatal period among AA women with Cronbach’s alpha scores ranging from 0.83-0.89 (Dolbier et al., 2013; Floyd James et al., 2023). The online survey also contained some author-developed question about prepartum and postpartum depression – “Have you ever had a diagnosis of depression or anxiety prior to pregnancy?” (Yes/No), “Were you ever depressed at any time after the birth of your child?” (Yes/No), Did having an unmet breastfeeding goal contribute to your feeling of depression?” (Yes/No).

### Data Analysis

#### Qualitative

Reflexive thematic analysis is a method involving inductive (and sometimes deductive) analysis of qualitative data incorporating researcher’s subjectivity to enrich descriptions and generate themes (Braun & Clarke, 2019, 2022). We chose this method because: 1) We used a qualitative descriptive design (Giorgi, 1992; Neergaard et al., 2009; Sandelowski, 2000) which makes our data suitable for reflexive thematic analysis, 2) TA has experiential knowledge of BG, and reflexive thematic analysis allows weaving in TA’s viewpoints into the analysis to generate rigorous findings that are rich in nuances and meanings (Braun & Clarke, 2019, 2022), and 3) We want to be transparent about how TA’s experiential knowledge of BG shaped data interpretation. Of note, themes were not generated prior to interviews as we conscientiously followed the six steps of reflexive thematic analysis identified by Braun and Clark (Braun & Clarke, 2021, 2022). *Data familiarization:* Transcripts of semi-structured interviews were downloaded from Zoom and edited by TA and CM to ensure that data were transcribed correctly (Byrne, 2022; Clark & Braun, 2013). TA read the transcripts twice to gain familiarization with the data after which transcripts were uploaded in Dedoose (dedoose, n.d.) for coding. *Systematic data coding:* Coding and development of themes were done by TA only, consistent with recommendations of quality reflexive thematic analysis (Braun & Clarke, 2019, 2021, 2022). Semantic and latent coding were used to identify obvious and implicit meanings of BG in the data (Braun & Clarke, 2019, 2021). Inductive and deductive approaches were used (Braun & Clarke, 2019; Clark & Braun, 2013). The inductive approach (dominant approach (Braun & Clarke, 2012)), allowed for open coding of data based on TA’s interpretation of patterns of the meaning of BG in the data (Braun & Clarke, 2019; Byrne, 2022; Clark & Braun, 2013); and codes were deductively reviewed to ensure they are valid to generate themes relevant to research question 1. *Generating initial themes:* Some codes with shared underlying meanings (e.g., codes about breastfeeding goals and motivation for setting those goals) were merged to generate initial themes. The qualitative research question examined participants’ experiences of BG; therefore, experiential orientation (Braun & Clarke, 2022; Byrne, 2022) (focused on participants’ meaning of BG) was used to interpret and organize codes into meaning-based interpretive stories (themes) about BG (Braun & Clarke, 2019, 2021). *Developing and reviewing themes:* Five initial themes were reviewed to determine if the codes that informed themes and subthemes fit well with the data, and if the themes are representative of the data and provide a comprehensive narrative of BG. One of the themes – prenatal expectations – was removed as it fits better as a subtheme (Braun & Clarke, 2012, 2021). CM reviewed the codes, themes, and data interpretations and affirmed that themes are representative of the data. *Refining, defining, and naming themes:* The final four themes were refined to ensure that each theme uniquely contributes to the research question and that all themes coherently provide a narrative about BG. Themes were also defined and named to provide meaning-based interpretive stories about BG rather than topic/domain summaries (Braun & Clarke, 2019, 2023). *Writing the report:* Final themes were arranged to provide a cogent narrative of BG (Braun & Clarke, 2012; Byrne, 2022), and a thematic map of BG was provided in the results (Figure 2).

**Figure 2.**
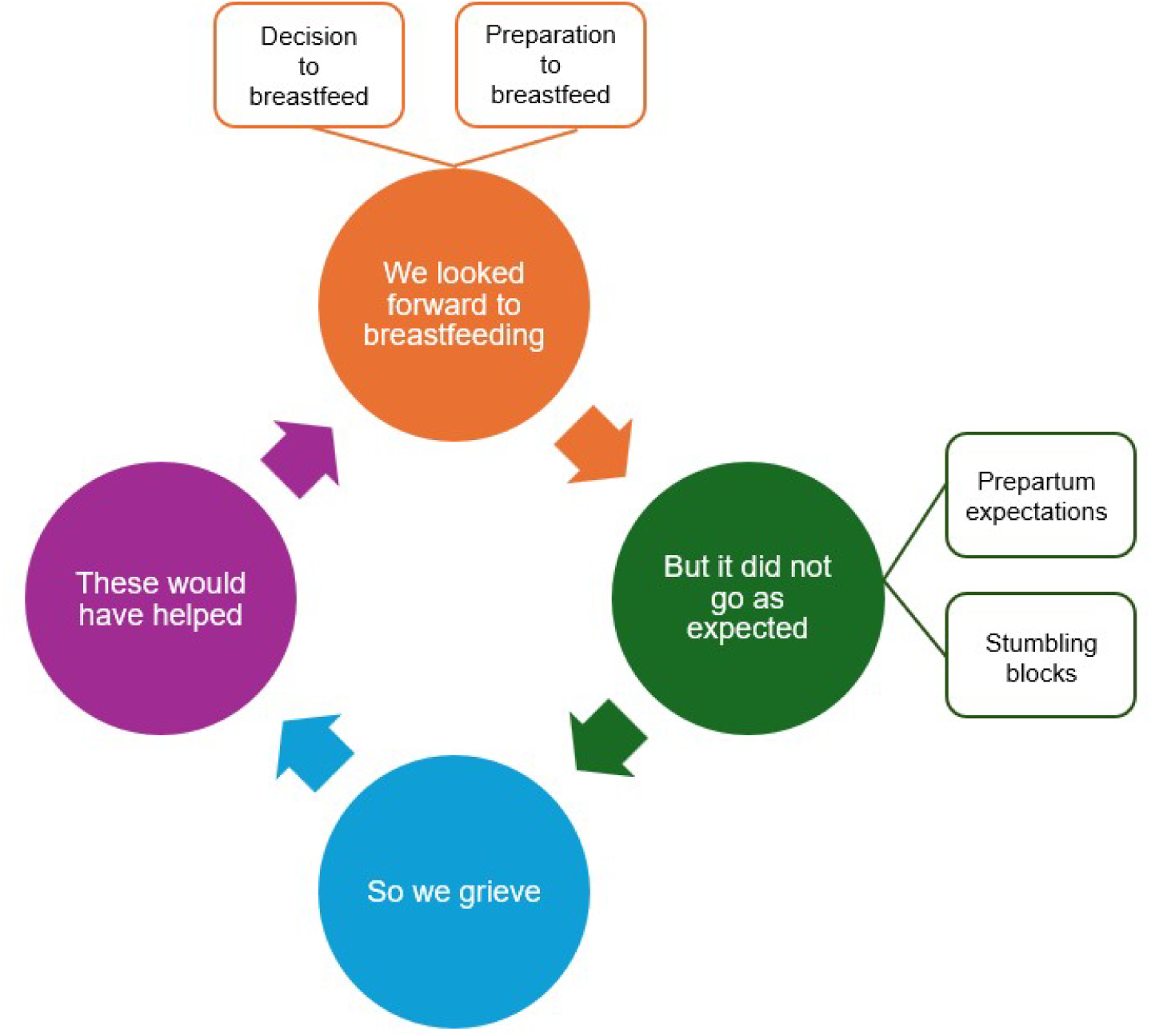
Thematic Map of Breastfeeding Grief.

#### Quantitative

Descriptive statistics were provided for EPDS scores and other quantitative variables. Each item on the EPDS was allotted a score between 0 (Never/No, not at all) to 3 (Yes, quite a lot/Yes, most of the time) (Cox et al., 1987). Participants with a scale score of 11 or higher were categorized as having depressive symptoms because a cut off score of 11 maximizes combined sensitivity and specificity of the EPDS (Levis et al., 2020). The sensitivity and specificity of the EPDS at 11-point cut-off are 81% and 88% respectively (Levis et al., 2020). Frequencies and percentages of participants with unmet breastfeeding goals who have PD symptoms were reported.

#### Data Integration

Consistent with convergent parallel design, qualitative and quantitative results were integrated following guidelines from Creswell and Plano Clark (2011) (Creswell & Plano Clark, 2011). Joint display of data (results), and side-by-side comparison (discussion) were used to integrate and interpret qualitative and quantitative results about the contributions of BG to PD symptoms.

### Rigor

We followed Creswell and colleagues (2018)’s guidelines for designing a mixed methods study (Creswell & Creswell, 2018). The EPDS tool is valid and reliable to screen for depressive symptoms in AA women. Maintenance of rigor in the qualitative strand was reported in our previous article.

## Results

### Quantitative Results

#### Demographic and Medical Characteristics

Participants’ mean age was 29.5 years (range 21-39 years). Participants were from 9 states across the United States. The majority of participants were married, employed and had completed a 4-year college education (Table 1). In addition, most participants were primiparous with no prior breastfeeding experience. (Table 2). Almost 88% of participants reported having a history of PD symptoms. Current PD symptoms examined using EPDS revealed a mean score of 12.88, with 9 participants having EPDS scores between 1-10 and 7 participants having EPDS scores >11. All participants reported that BG, as a result of an unmet breastfeeding goal, contributed to their PD symptoms (Table 2).

#### Qualitative Results

Meaning-based themes in reflexive thematic analysis capture the meaning of the phenomenon investigated and tells an interpretive story about that meaning (Braun & Clarke, 2019, 2023).

Four meaning-based themes about BG (two having sub-themes) were generated; these include *We looked forward to breastfeeding, But it did not go as expected*, *So we grieve*, and *These would have helped*.

#### We looked forward to breastfeeding

Participants narrated how they longed for and prepared to breastfeed. This theme was further divided into two subthemes – a) decision to breastfeed, and b) preparation to breastfeed.

##### Decision to breastfeed

Participants’ goals were to partially breastfeed and/or to exclusively breastfeed for a certain duration and most participants made these decisions before getting pregnant:

> “It was a goal I had set before my pregnancy, so it would have been really nice for me to you know, achieve it.” [P9]

Additionally, participants identified various reasons for setting those goals. These include knowledge about the health benefits of breastfeeding obtained from personal research, friends, or health professionals; love for child; familial norm; and breastfeeding being a natural method of infant feeding:

> “My breastfeeding goal was something I set because I love my baby, so I want to give her something really good. I heard from the experience, from all the models, you know, research online. I heard other mothers’ reviews. Oh, this this really helps my child.” [P3]

> “It was kind of cause of the doctors, and everybody telling me to breastfeed, and also just because how convenient it is as well, you know, you don’t have to get up and prep a bottle, make sure it’s warm, and don’t have to do none of it.” [P8]

> “I have a younger brother who is around 3-6 years, and I watched him being breastfed until 2 years.” [P15]

##### Preparation to breastfeed

Participants’ enthusiasm to breastfeed reflected in their preparations:

> “Prior to me giving birth, I also had a breast pump. So, I also um experimented with that just to make sure that I, you know, like I could do this, and I knew what I was doing before like the baby came.” [P12]

> “I already knew I had to breastfeed the baby because you know, I had been going for you know checkups, I had been having, you know I, I had been educated about breastfeeding.” [P9]

#### But it did not go as expected

Despite having high hopes and preparing to breastfeed, participants did not meet their breastfeeding goals. This theme was further divided into two subthemes – a) prepartum expectations, and b) stumbling blocks.

##### Prepartum expectations

During pregnancy, participants had positive predictions about their breastfeeding experiences:

> “I never thought like I’ll have an issue with. I was just told that, you know, “when the baby comes, the breast milk will come, I, like it, it happens automatically. It’s just biology!””. [P13]

##### Stumbling blocks

This subtheme had the highest number of excerpts compared to other themes and subthemes. Participants encountered several challenges that prevented them from meeting their breastfeeding goals. These include prepartum and postpartum depression, postpartum frustration and anxiety, perceived insufficient milk, breast and nipple pain, poor latch, sleep deprivation, stress, tongue-tie, lack of/inadequate support from family and health professionals, mastitis, financial constraints, time management, maternal illness, too much pressure to breastfeed, and experience of racism:

> “I think the main reason is postpartum depression and just being overwhelmed since I was a first-time mom, and I had expectations.” [P17]

> “One day I started feeling pain in my right nipple, then the pain just kept on increasing, and I started having these flu-like symptoms. And then I had to visit my doctor and at first, she said that that’s normal for first-time moms, but the pain did not stop. So, after I think 1 week it got worse. I was vomiting. Yeah, and all those signs. Then I went and that’s when I was not that I have mastitis, so this right breast blocked completely.” [P15]

> “I feel like even the doctors feel like, maybe some of us from a certain racial group are not that serious when they talk about breastfeeding.” [P12]

#### So we grieve

##### Participants experienced BG as sadness and depression

> “It was so depressing because I started to feel like I’m not developing this…okay. I was saying that at some point I started feeling like I’m being disconnected with my son because when I was breastfeeding him, there was that I know you understand that feeling between a mother and a child but when that stopped, I felt the disconnection starting to grow, yeah and I started feeling I started detaching from friends, from family. Yeah, it was not a good experience.” [P15]

> “I’m too low, like, like, I’m depressed of those low moments.” [P14]

#### These would have helped

In the final theme, participants outlined the resources they think would have helped them to achieve their breastfeeding goals including adequate support from spouses, other family members, and health professionals; and breastfeeding education for women and their spouses/partners. Participants prefer breastfeeding education comprising possible breastfeeding challenges, hands-on training on how to breastfeed, access to lactation consultants in the outpatient postpartum period, and detailed information about their services, and access to a doula:

> “I feel like there should be someone (a lactation consultant) coming home with you.” [P12]

> “Um I wish that they were like uh I wish I was told, you know, when I was pregnant, that I could face issues like this, and maybe a possible solution to like avoid things like this. If I had access to that, I think it would have like helped me or like prepared me, I think it um it just came as a shock.” [P5]

> “Um I wish that I had been more informed, maybe from my pediatrician or from my gynecologist, the doctor who delivered my baby, um from the hospital on who or how I could get assistance, because I know that there are lactation consultants, but I also am not familiar with how do they bill insurance? Is it free? Do I pay? How much is it?

> Frequency? I didn’t know, and they didn’t share that.” [P19]

#### Mixed Methods results

From the qualitative results, women with BG reported experiencing PD symptoms. This finding is congruent with the quantitative results where participants also reported having a history of PD symptoms (by answering *yes* to the question about ‘ever depressed postpartum’) and current PD symptoms (almost 44% having EPDS score >11). Though not the primary focus of the study, 81.3% of participants received breastfeeding education, either during pregnancy or the inpatient postpartum period (quantitative results), which converges with *preparation to breastfeed* subtheme in the qualitative results. Similarly, 87.5% of participants decided to breastfeed before giving birth (quantitative results) also corresponds with *decision to breastfeed* subtheme in the qualitative results. Therefore, according to Sandelowski et al. (2006) (Sandelowski et al., 2006), confirmation has occurred because the same findings about BG and PD symptoms are present in both qualitative and quantitative results (Table 3).

## Discussion

This exploratory study aimed to examine the possible contributions of BG to PD symptoms using both qualitative and quantitative data. Though the EPDS has been consistently used to screen women for PD in the first year postpartum (Míguez & Vázquez, 2023; New York State Office of Mental Health & New York State Department of Health, 2023; Robbins et al., 2023), evidence suggest that PD symptoms may persist until 11 years postpartum (Netsi et al., 2018). Using side-by-side comparison, we cautiously interpreted qualitative and quantitative findings. *Quantitative:* All participants indicated that their experience of BG was a result of unmet breastfeeding goals, and it contributed to their past/current PD symptoms. This was further revealed by almost 44% of participants having EPDS score >11, which may indicate the presence of underlying depression/mood disorders. *Qualitative:* In the ‘*So we grieve*’ theme focused on BG; participants expressed that they were depressed because they were unable to meet their breastfeeding goals. *Interpretation:* Our combined survey responses and qualitative data suggests that women experiencing BG may be likely to have PD symptoms. Women may experience BG because breastfeeding, an act performed by biological women (Williamson et al., 2023) is a behavioral component of maternal identity (Perun, 2013; Sen, 2022). Notably, depression is one of Kubler-Ross’ stages of grief (Kübler-Ross, 1969), hence our findings correlate with Kubler-Ross’ grief model.

Breastfeeding and PD have a bidirectional relationship (Figueiredo et al., 2014). Women with PD symptoms have shorter breastfeeding duration compared to those without PD symptoms (Bascom & Napolitano, 2016). Accordingly, women who breastfeed exclusively for >3 months had lower PD symptoms compared to those who did not (Figueiredo et al., 2014). One study with a similar research focus, reported reduced PD symptoms among women who met their prepartum breastfeeding exclusivity goals (E. F. Gregory et al., 2015). Breastfeeding’s protective effect on maternal mental health (Tucker & O’Malley, 2022) is attributed to its neuroendocrine responses to stress via the release of lactation hormones – oxytocin and prolactin – which improve mood (Pope & Mazmanian, 2016). Oxytocin, the love hormone, is responsible for milk ejection, and it also decreases cortisol secretion to reduce stress (Viero et al., 2010). Prolactin, the adaptive hormone, functions in milk synthesis and modulation of stress responses during breastfeeding (Faron-Górecka et al., 2023).

Our secondary findings were also interpreted. *Quantitative:* The majority of participants decided to breastfeed before the birth of their infants and also received breastfeeding education during pregnancy or in the inpatient postpartum period. *Qualitative:* In the subthemes ‘*decision to breastfeed*’ and ‘*preparation to breastfeed*’, participants described when they decided to breastfeed and what motivated their decisions; and their preparations to breastfeed. *Interpretation:* Participants were enthusiastic about breastfeeding but were unable to meet their breastfeeding goals, which may have contributed to their BG and PD symptoms. Authors of previous studies reported the mediating effect of intention on the inverse relationship between breastfeeding and PD (Borra et al., 2015; Rosenbaum et al., 2020). Among women who were not depressed during pregnancy, those who had planned to breastfeed but could not had higher risks for PD compared to those who had no intention to breastfeed (Borra et al., 2015; Rosenbaum et al., 2020).

Two qualitative themes (‘*but it did not go as expected*’ and ‘*these would have helped*’) with no corresponding quantitative data were not included in the mixed methods interpretation. In the ‘*but it did not go as expected*’ theme, participants narrated several challenges that prevented them from meeting their breastfeeding goals. Consistent with earlier studies (Author & Lucas, 2022; Author et al., 2024; Author et al., 2023; Gianni et al., 2019), perceived insufficient milk and breast and nipple pain continue to be leading reasons why AA women stop breastfeeding earlier than desired. Participants also listed resources they think would have helped them to meet their breastfeeding goals in the ‘*these would have helped*’ theme. Similar findings were reported in previous studies (Author et al., 2025; Spencer et al., 2015).

As this is an exploratory/pilot study, future studies with sufficient power and sample size are needed to examine the correlation between BG and PD/PD symptoms. Additionally, it is important to examine BG in other populations. Finally, breastfeeding interventions should target the barriers identified in the ‘*stumbling blocks*’ subtheme and resources listed in the ‘*these would have helped*’ theme.

### Strengths and Limitations

This study has unique strengths and some limitations. First, the triangulation of qualitative and qualitative data to understand BG and its possible role in PD symptoms is novel. This method provided a more comprehensive overview of BG and PD symptoms compared to using qualitative or quantitative data only. Second, the relationship between BG and PD symptoms has not been examined quantitatively, our findings provide a hypothesis/reference for further examination of these variables. Lastly, the use of reflexive thematic analysis facilitated the weaving in of TA’s viewpoints into the qualitative analysis to generate rigorous findings that are rich in nuances and meanings. The limitations of the study include low sample size to conduct a correlational analysis between BG and PD symptoms. As with internet-based recruitment studies, the population is skewed to participants with higher income and education.

Confirming this, the majority of participants (62.6%) were employed. Remarkedly, the EPDS was designed to screen for PD during the first year postpartum hence, the relevance of the EPDS scores to women with children older than 1 year is limited. Considering recent reports of prolonged PD lasting longer than 1 year postpartum (Netsi et al., 2018; Putnick et al., 2020), it is important to develop screening tools to measure PD in women after 1 year postpartum. In addition, future studies to examine if low-income AA women have more breastfeeding challenges compared to high-income women and if this impact their experience of BG are warranted.

### Implications for Practice and/or Policy

Our findings may have implications for clinical practice and policy. There is a need for increased awareness and detailed information about the services of lactation consultants, as participants wished they knew their ‘modus operandi’. The majority of participants decided to breastfeed because of their knowledge of the benefits of breastfeeding. It is important for clinicians to continue to offer breastfeeding education to women during pregnancy and incorporate possible breastfeeding challenges into the education. Also, screening and providing mental health support for women experiencing BG may help to prevent PD. Finally, policies extending the duration of paid maternity leave to at least 6 months, and policies to reduce systemic racism are warranted to promote breastfeeding and potentially circumvent BG and PD.

## Conclusions

Postpartum depression continues to be the leading mental health complication that women experience after childbirth. This study elucidates the possible contributions of BG to PD symptoms. Hence, examining women’s experiences of BG is crucial for understanding and mitigating PD risk factors during the critical postpartum period. Importantly, as our sample comprised women with children aged 2 years, continuous screening of women for PD until 2 years postpartum may be clinically valuable.

## Data Availability Statement

To protect participant privacy, the datasets presented in this article are prohibited from being made public. Nonetheless, a reasonable request to access the datasets should be made to the corresponding author.

## CRediT authorship contribution statement

**TA:** Conceptualization; Methodology; Investigation, Validation, Data collection, Data curation, Formal analysis, Writing – original draft, Writing – review and editing. **CM:** Formal analysis, Validation, Visualization, Writing – review and editing. **RK**: Formal analysis, Validation, Visualization, Writing – review and editing. **BO:** Data curation. **SS:** Supervision, Writing – review and editing**. GL**: Supervision, Writing – review and editing.

## Supplementary Material

Additional supplementary materials including the reflexivity process throughout the study (Supplement 1), Creswell and Creswell (2018) mixed methods research checklist (Supplement 2), and additional participant quotes (Supplement 3) are included.

## Data Availability

All data produced in the present study are available upon reasonable request to the corresponding author

## Acknowledgements

Research reported in this publication was supported by the Burton Post-Doctoral Fellowship at the University of Utah College of Nursing. The content is solely the responsibility of the authors and does not necessarily represent the official views of the University of Utah. TA acknowledges Professor Cheryl Beck at the University of Connecticut for her great teaching on mixed methods research. Tumilara Aderibigbe had full access to all the data in the study and takes responsibility for the integrity of the data and the accuracy of the data analysis.

## References

1. Author, O., & Lucas, R. (2022). Exclusive breastfeeding in African American women: A concept analysis. Journal of Advanced Nursing, 79(5), 1699–1713. 10.1111/jan.15301

2. Author, T. (2026). It is mentally challenging – Understanding breastfeeding grief during the postpartum period: A qualitative descriptive study. Women and Birth, 39(1).

3. Author, T., Adeleye, K., Author, S., & Author, G. (2025). A Narrative Review of Culturally Informed Breastfeeding Interventions for African American Women. Journal of Perinatal and Neonatal Nursing, 39(2), 137–149. 10.1097/JPN.0000000000000912

4. Author, T., Kelleher, S., Henderson, W., Prescott, S., Young, E., & Lucas, R. (2024). COMT Variants are Associated With Breast and Nipple Pain. The Journal of Pain, 25(9), 104568. 10.1016/j.jpain.2024.104568

5. Author, T., Kent-Marvick, J., Austin, S., Macias, S. N., Simonsen, S. E., Crandall, L., Ellis, J. A., Maughan, M. E., Madrigal, C., Ward, R., & Taylor-Swanson, L. (2026). Waning Moon: Revising a Culturally Informed Intervention with Perimenopausal American Indian and Alaska Native Women. EXPLORE, 103347. 10.1016/j.explore.2026.103347

6. Author, T., Srisopa, P., Henderson, W., & Lucas, R. (2023). Meta- ethnography on the Experiences of Women From Around the World Who Exclusively Breastfed Their Full-term Infants. *Journal of Obstetrics*, Gynecology, and Neonatal Nursing, 53(2), 120–131. 10.1016/j.jogn.2023.11.008

7. Author, T., Walsh, S., Henderson, W. A., & Lucas, R. (2023). Psychometric testing of the breastfeeding self-efficacy scale to measure exclusive breastfeeding in African American women: A cross-sectional study. Frontiers in Public Health, 11(1196510).

8. Aggarwal, R., & Ranganathan, P. (2019). Study designs: Part 2 - Descriptive studies. In Perspectives in Clinical Research (Vol. 10, Number 1, pp. 34–36). Wolters Kluwer Medknow Publications. 10.4103/picr.PICR_154_18

9. Bascom, E. M. E., & Napolitano, M. A. (2016). Breastfeeding Duration and Primary Reasons for Breastfeeding Cessation among Women with Postpartum Depressive Symptoms. Journal of Human Lactation, 32(2), 282–291. 10.1177/0890334415619908

10. Borra, C., Iacovou, M., & Sevilla, A. (2015). New Evidence on Breastfeeding and Postpartum Depression: The Importance of Understanding Women’s Intentions. Maternal and Child Health Journal, 19(4), 897–907. 10.1007/s10995-014-1591-z

11. Braun, V., & Clarke, V. (2012). Thematic analysis. In APA handbook of research methods in psychology, Vol 2: Research designs: Quantitative, qualitative, neuropsychological, and biological. (pp. 57–71). American Psychological Association. 10.1037/13620-004

12. Braun, V., & Clarke, V. (2019). Reflecting on reflexive thematic analysis. *Qualitative Research in Sport*, Exercise and Health, 11(4), 589–597. 10.1080/2159676X.2019.1628806

13. Braun, V., & Clarke, V. (2021). One size fits all? What counts as quality practice in (reflexive) thematic analysis? Qualitative Research in Psychology, 18(3), 328–352. 10.1080/14780887.2020.1769238

14. Braun, V., & Clarke, V. (2022). Thematic Analysis: A Practical Guide (1st ed.). SAGE.

15. Braun, V., & Clarke, V. (2023). Toward good practice in thematic analysis: Avoiding common problems and be(com)ing a knowing researcher. International Journal of Transgender Health, 24(1), 1–6. 10.1080/26895269.2022.2129597

16. Brown, A. (2018). What Do Women Lose if They Are Prevented From Meeting Their Breastfeeding Goals? Clinical Lactation, 9(4). 10.1891/2158

17. Brown, A. (2019). Why Breastfeeding Grief and Trauma Matter. Pinter & Martin.

18. Byrne, D. (2022). A worked example of Braun and Clarke’s approach to reflexive thematic analysis. Quality and Quantity, 56(3), 1391–1412. 10.1007/s11135-021-01182-y

19. Cannon, C., & Nasrallah, H. (2019). A Focus on Postpartum Depression among African American Women: A Literature Review. Annals of Clinical Psychiatry, 31(2). 10.1177/104012371903100206

20. Ceballos, M., Wallace, G., & Goodwin, G. (2017). Postpartum Depression among African-American and Latina Mothers Living in Small Cities, Towns, and Rural Communities. Journal of Racial and Ethnic Health Disparities, 4(5), 916–927. 10.1007/s40615-016-0295-z

21. Centers for Disease Control and Prevention. (2022). Breastfeeding Report Card, United States, 2022. https://www.cdc.gov/breastfeeding/data/reportcard.htm

22. Cherryholmes, C. H. (1992). Notes on Pragmatism and Scientific Realism. Educational Researcher, 21(6), 13–17.

23. Clark, V., & Braun, V. (2013). Successful Qualitative Research: A practical guide for beginners (1st ed.). SAGE Publications.

24. Cox, J. L., Holden, J. M., & Sagovsky, R. (1987). Detection of Postnatal Depression: Development of the 10-item Edinburgh Postnatal Depression scale. British Journal of Psychiatry, 150(JUNE), 782–786. 10.1192/bjp.150.6.782

25. Creswell, J. W. (1999). Mixed-Method Research: Introduction and Application. In J. C. Gregory (Ed.), Handbook of Educational Policy (1st ed., 18). Academic Press.

26. Creswell, J. W., & Plano Clark, V. L. (2011). Designing and Conducting Mixed Methods Research (2nd ed.). SAGE Publications Inc.

27. Creswell J. W., & Creswell, J. D. (2018). Research Design: Qualitative, Quantitative and Mixed Methods Approaches (5th ed.). SAGE Publications Inc.

28. dedoose. (n.d.). *dedoose: Great Research Made Easy*. Retrieved January 6, 2024, from https://www.dedoose.com/

29. Demirci, J. (2022). Breastfeeding Grief. The Journal of Perinatal & Neonatal Nursing, 36(2), 115–117. 10.1097/JPN.0000000000000650

30. Dolbier, C. L., Rush, T. E., Sahadeo, L. S., Shaffer, M. L., & Thorp, J. (2013). Relationships of race and socioeconomic status to postpartum depressive symptoms in rural African American and non-hispanic white women. Maternal and Child Health Journal, 17(7), 1277–1287. 10.1007/s10995-012-1123-7

31. Doyle, L., McCabe, C., Keogh, B., Brady, A., & McCann, M. (2020). An overview of the qualitative descriptive design within nursing research. Journal of Research in Nursing, 25(5), 443–455. 10.1177/1744987119880234

32. Faron-Górecka, A., Latocha, K., Pabian, P., Kolasa, M., Sobczyk-Krupiarz, I., & Dziedzicka-Wasylewska, M. (2023). The Involvement of Prolactin in Stress-Related Disorders. In International Journal of Environmental Research and Public Health (Vol. 20, Number 4). MDPI. 10.3390/ijerph20043257

33. Figueiredo, B., Canário, C., & Field, T. (2014). Breastfeeding is negatively affected by prenatal depression and reduces postpartum depression. Psychological Medicine, 44(5), 927–936. 10.1017/S0033291713001530

34. Finlay, L. (2002). Negotiating the swamp: The opportunity and challenge of reflexivity in research practice. Qualitative Research, 2(2), 209–230. 10.1177/146879410200200205

35. Floyd James, K., Smith, B. E., Robinson, M. N., Thomas Tobin, C. S., Bulles, K. F., & Barkin, J. L. (2023). Factors Associated with Postpartum Maternal Functioning in Black Women: A Secondary Analysis. Journal of Clinical Medicine, 12(2). 10.3390/jcm12020647

36. Getahun, D., Oyelese, Y., Peltier, M., Yeh, M., Chiu, V. Y., Takhar, H., Khadka, N., Mensah, N., Avila, C., & Fassett, M. J. (2023). Trends in Postpartum Depression by Race/Ethnicity and Pre-pregnancy Body Mass Index. American Journal of Obstetrics and Gynecology, 228(1), S122–S123. 10.1016/j.ajog.2022.11.248

37. Gianni, M. L., Bettinelli, M. E., Manfra, P., Sorrentino, G., Bezze, E., Plevani, L., Cavallaro, G., Raffaeli, G., Crippa, B. L., Colombo, L., Morniroli, D., Liotto, N., Roggero, P., Villamor, E., Marchisio, P., & Mosca, F. (2019). Breastfeeding difficulties and risk for early breastfeeding cessation. Nutrients, 11(10). 10.3390/nu11102266

38. Giorgi, A. (1992). Description versus Interpretation: Competing Alternative Strategies for Qualitative Research. Journal of Phenomenological Psychology, 23(2), 119–135. 10.1163/156916292X00090

39. Gregory, E. F., Butz, A. M., Ghazarian, S. R., Gross, S. M., & Johnson, S. B. (2015). Are unmet breastfeeding expectations associated with maternal depressive symptoms? Academic Pediatrics, 15(3), 319–325. 10.1016/j.acap.2014.12.003

40. Gribble, K. D., Bewley, S., & Dahlen, H. G. (2023). Breastfeeding grief after chest masculinisation mastectomy and detransition: A case report with lessons about unanticipated harm. Frontiers in Global Women’s Health, 4. 10.3389/fgwh.2023.1073053

41. Hennink, M., & Kaiser, B. N. (2022). Sample sizes for saturation in qualitative research: A systematic review of empirical tests. Social Science and Medicine, 292. 10.1016/j.socscimed.2021.114523

42. Howell, E. A., Mora, P. A., Horowitz, C. R., & Leventhal, H. (2005). Racial and ethnic differences in factors associated with early postpartum depressive symptoms. Obstetrics and Gynecology, 105(6), 1442–1450. 10.1097/01.AOG.0000164050.34126.37

43. Kruger, L. J., Rodgers, R. F., Long, S. J., & Lowy, A. S. (2019). Individual interviews or focus groups? Interview format and women’s self-disclosure. International Journal of Social Research Methodology, 22(3), 245–255. 10.1080/13645579.2018.1518857

44. Kübler-Ross, E. (1969). On Death and Dying (1st ed.). Routledge. 10.4324/9780203010495

45. Latendresse, G., Pentecost, R., Iacob, E., Simonsen, S., Williams, M., Thompson, N., & Hogue, C. (2023). A group telehealth intervention for rural perinatal depression and anxiety: A pilot study. Journal of Rural Mental Health, 47(1), 20–29. 10.1037/rmh0000213

46. Levis, B., Negeri, Z., Sun, Y., Benedetti, A., & Thombs, B. D. (2020). Accuracy of the Edinburgh Postnatal Depression Scale (EPDS) for screening to detect major depression among pregnant and postpartum women: Systematic review and meta-analysis of individual participant data. The BMJ, 371. 10.1136/bmj.m4022

47. Liu, C. H., & Tronick, E. (2013). Rates and predictors of postpartum depression by race and ethnicity: Results from the 2004 to 2007 New York city PRAMS survey (pregnancy risk assessment monitoring system). Maternal and Child Health Journal, 17(9), 1599–1610. 10.1007/s10995-012-1171-z

48. Míguez, M. C., & Vázquez, M. B. (2023). Prevalence of postpartum major depression and depressive symptoms in Spanish women: A longitudinal study up to 1 year postpartum. Midwifery, 126. 10.1016/j.midw.2023.103808

49. Morgan, D. L. (2007). Paradigms Lost and Pragmatism Regained: Methodological Implications of Combining Qualitative and Quantitative Methods. Journal of Mixed Methods Research, 1(1), 48–76. 10.1177/2345678906292462

50. Morse, J. M. (1991). Approaches to qualitative–quantitative methodological triangulation. Nursing Research, 40(1), 120–123. 10.1017/S0714980800008576

51. Neergaard, M. A., Olesen, F., Andersen, R. S., & Sondergaard, J. (2009). Qualitative description-the poor cousin of health research? In BMC Medical Research Methodology (Vol. 9, Number 1). 10.1186/1471-2288-9-52

52. Netsi, E., Pearson, R. M., Murray, L., Cooper, P., Craske, M. G., & Stein, A. (2018). Association of persistent and severe postnatal depression with child outcomes. JAMA Psychiatry, 75(3), 247–253. 10.1001/jamapsychiatry.2017.4363

53. New York State Office of Mental Health, & New York State Department of Health. (2023). Postpartum Depression Screening Protocols and Tools: A Review of Evidence on Adequacy and Equity. https://www.postpartum.net/learn-more/anxiety/

54. Onyewuenyi, T. L., Peterman, K., Zaritsky, E., Ritterman Weintraub, M. L., Pettway, B. L., Quesenberry, C. P., Nance, N., Surmava, A. M., & Avalos, L. A. (2023). Neighborhood Disadvantage, Race and Ethnicity, and Postpartum Depression. JAMA Network Open, E2342398. 10.1001/jamanetworkopen.2023.42398

55. Patton, M. Q. (1990). Qualitative evaluation and research methods (2nd ed.). SAGE Publications Inc.

56. Perun, M. B. (2013). Maternal identity of women in the postpartum period. Journal of Education Culture and Society, 1(2013). 10.15503/jecs20131-95-105

57. Pope, C. J., & Mazmanian, D. (2016). Breastfeeding and postpartum depression: An overview and methodological recommendations for future research. In Depression Research and Treatment (Vol. 2016). Hindawi Publishing Corporation. 10.1155/2016/4765310

58. Putnick, D. L., Sundaram, R., Bell, E. M., Ghassabian, A., Goldstein, R. B., Robinson, S. L., Vafai, Y., Gilman, S. E., & Yeung, E. (2020). Trajectories of Maternal Postpartum Depressive Symptoms. Pediatrics, 146(5). 10.1542/peds.2020-0857

59. Robbins, C. L., Ko, J. Y., D’Angelo, D. V., von Essen, B. S., Bish, C. L., Kroelinger, C. D., Tevendale, H. D., Warner, L., & Barfield, W. (2023). Timing of Postpartum Depressive Symptoms. Preventing Chronic Disease, 20. 10.5888/PCD20.230107

60. Robinson, C. (2018). Misshapen motherhood: Placing breastfeeding distress. *Emotion*, Space and Society, 26, 41–48. 10.1016/j.emospa.2016.09.008

61. Rosenbaum, D. L., Gillen, M. M., & Markey, C. H. (2020). Feeling let down: An investigation of breastfeeding expectations, appreciation of body functionality, self-compassion, and depression symptoms. Appetite, 154. 10.1016/j.appet.2020.104756

62. Sandelowski, M. (2000). Focus on Research Methods Whatever Happened to Qualitative Description? Research in Nursing & Health, 23.

63. Sandelowski, M. (2010). What’s in a name? Qualitative description revisited. Research in Nursing and Health, 33(1), 77–84. 10.1002/nur.20362

64. Sandelowski, M., Voils, C. I., & Barroso, J. (2006). Defining and Designing Mixed Research Synthesis Studies. Research in the SchoolsJ: A Nationally Refereed Journal Sponsored by the Mid-South Educational Research Association and the University of Alabama, 13(1), 29.

65. Sen, S. (2022). Breast Milk and Breastfeeding: Benefits, Barriers, Maternal Predictors, and Opportunities for Innovation. Clinical Therapeutics, 44(2), 170–171. 10.1016/j.clinthera.2021.11.004

66. Spencer, B., Wambach, K., & Domain, E. W. (2015). African American Women’s Breastfeeding Experiences: Cultural, Personal, and Political Voices. Qualitative Health Research, 25(7), 974–987. 10.1177/1049732314554097

67. Stanhope, K. K., Perreira, K. M., Isasi, C. R., LeCroy, M. N., Daviglus, M. L., Gonzalez, F., Gallo, L. C., Poelinz, C. M., & Suglia, S. F. (2024). Differences in Breastfeeding Initiation and Duration by Migration History in the Hispanic Community Health Study/Study of Latinos. Breastfeeding Medicine, 19(12), 957–963. 10.1089/bfm.2024.0162

68. Tabb, K. M., Hsieh, W. J., Gavin, A. R., Eigbike, M., Faisal-Cury, A., Hajaraih, S. K. M., Huang, W. hao D., Laurent, H., Carter, D., Nidey, N., Ryckman, K., & Zivin, K. (2020). Racial differences in immediate postpartum depression and suicidal ideation among women in a Midwestern delivery hospital. Journal of Affective Disorders Reports, 1. 10.1016/j.jadr.2020.100008

69. Tashakkori, A., & Creswell, J. W. (2007). Editorial: Exploring the Nature of Research Questions in Mixed Methods Research. In Journal of Mixed Methods Research (Vol. 1, Number 3, pp. 207–211). 10.1177/1558689807302814

70. Trainor, L. R., & Bundon, A. (2021). Developing the craft: reflexive accounts of doing reflexive thematic analysis. *Qualitative Research in Sport*, Exercise and Health, 13(5), 705–726. 10.1080/2159676X.2020.1840423

71. Tucker, Z., & O’Malley, C. (2022). Mental Health Benefits of Breastfeeding: A Literature Review. Cureus. 10.7759/cureus.29199 University of Utah. (2023). Ubox. https://box.utah.edu/

72. Viero, C., Shibuya, I., Kitamura, N., Verkhratsky, A., Fujihara, H., Katoh, A., Ueta, Y., Zingg, H. H., Chvatal, A., Sykova, E., & Dayanithi, G. (2010). Oxytocin: Crossing the bridge between basic science and pharmacotherapy. In CNS Neuroscience and Therapeutics (Vol. 16, Number 5). 10.1111/j.1755-5949.2010.00185.x

73. Williamson, T., Wagstaff, D. L., Goodwin, J., & Smith, N. (2023). Mothering Ideology: A Qualitative Exploration of Mothers’ Perceptions of Navigating Motherhood Pressures and Partner Relationships. Sex Roles, 88(1–2), 101–117. 10.1007/s11199-022-01345-7

74. World Health Organization. (2026). *Breastfeeding*. https://www.who.int/health-topics/breastfeeding#tab=tab_2

