## Supplementary material for "Preventing postpartum depression through mitigating breastfeeding grief: A convergent parallel mixed methods study": Tables

**Table 1 Demographic characteristics of participants (n=16)**

| **Variable** | **Frequency (N)** | **Percentage (%)** |
| --- | --- | --- |
| Age, *mean (SD)* | 29.50 (4.41) |  |
| Location |  |  |
| Arizona | 1 | 6.3 |
| California | 1 | 6.3 |
| Florida | 3 | 18.8 |
| Georgia | 2 | 12.2 |
| Indiana | 1 | 6.3 |
| New York | 3 | 18.8 |
| Pennsylvania | 1 | 6.3 |
| Texas | 3 | 18.8 |
| Washington | 1 | 6.3 |
| Education |  |  |
| High school diploma or GED | 1 | 6.3 |
| Some college | 2 | 12.5 |
| Graduated 2-year college | 4 | 25.0 |
| Graduated 4-year college | 8 | 50.0 |
| Master’s degree | 1 | 6.3 |
| Employment |  |  |
| Unemployed/house keeping | 3 | 18.8 |
| Student (unemployed) | 1 | 6.3 |
| Employed part-time | 5 | 31.3 |
| Employed full-time | 5 | 31.3 |
| Marital status |  |  |
| Married | 6 | 37.5 |
| Single (never married) | 5 | 31.3 |
| In a domestic partnership | 4 | 25.0 |
| Separated | 1 | 6.3 |

**Table 2** **Medical/Lactation characteristics of participants (n=16)**

| **Variable** | **Frequency (N)** | **Percentage (%)** |
| --- | --- | --- |
| EPDS, *mean (SD)* | 12.88 (7.99) | range: 1-26 |
| Age of child in months, *mean (SD)* | 13.56 (6.13) | range: 6-24 |
| Parity |  |  |
| Primipara | 11 | 68.8 |
| Multipara | 5 | 31.3 |
| Mode of delivery |  |  |
| Vaginal | 13 | 81.2 |
| Cesarean section | 3 | 18.8 |
| Diagnosis of depression/anxiety before pregnancy |  |  |
| Yes | 2 | 12.5 |
| No | 14 | 87.5 |
| Complications of pregnancy (e.g., gestational diabetes or hypertension, preeclampsia) |  |  |
| Yes | 2 | 12.5 |
| No | 14 | 87.5 |
| Decision to breastfeed infant was made |  |  |
| Before pregnancy | 8 | 50.0 |
| During pregnancy | 6 | 37.5 |
| After birth | 2 | 12.5 |
| Breastfeeding education during pregnancy or inpatient postpartum |  |  |
| Yes | 13 | 81.2 |
| No | 3 | 18.8 |
| Prior breastfeeding experience |  |  |
| Yes | 3 | 18.8 |
| No | 13 | 81.2 |
| Exclusive breastfeeding duration |  |  |
| 0 hour-1 week | 2 | 12.5 |
| 2-6 weeks | 3 | 18.8 |
| 7 weeks-3 months | 5 | 31.3 |
| 4-5 months | 4 | 25.0 |
| ≥6 months | 2 | 12.5 |
| Ever had postpartum depressive symptoms |  |  |
| Yes | 14 | 87.5 |
| No | 2 | 12.5 |
| Breastfeeding grief contributed to postpartum depression symptoms |  |  |
| Yes | 16 | 100.0 |

**Table 3** Joint display of findings on breastfeeding grief and postpartum depressive symptoms

| **Breastfeeding grief and postpartum depressive symptoms** | |
| --- | --- |
| Quantitative results | Qualitative results |
| 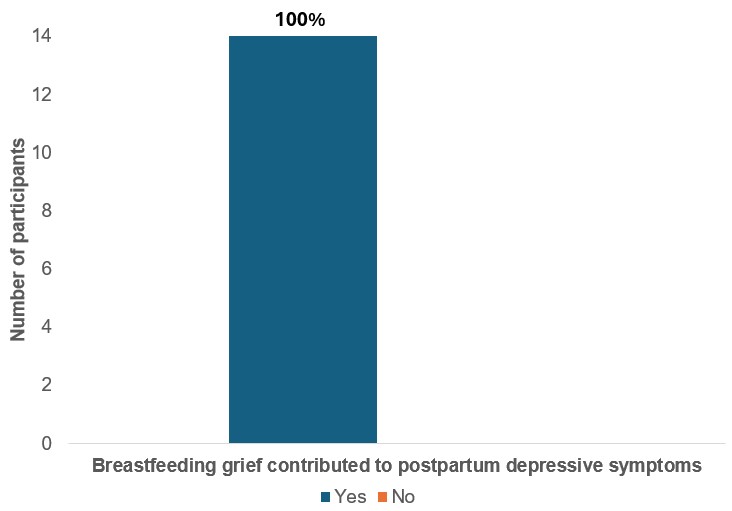 | Theme ‘*So we grieve*’  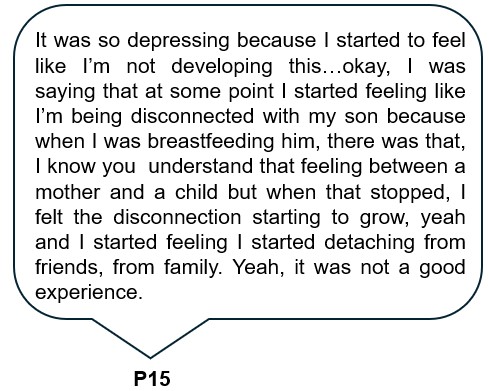 |
| 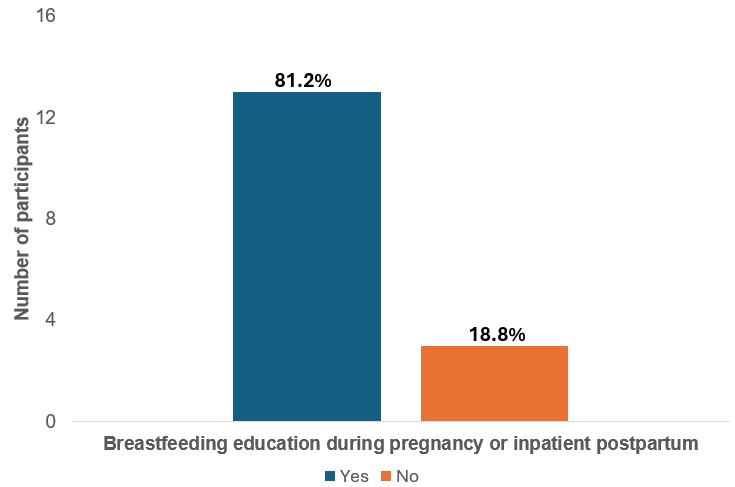 | Subtheme ‘*Preparation to breastfeed’*  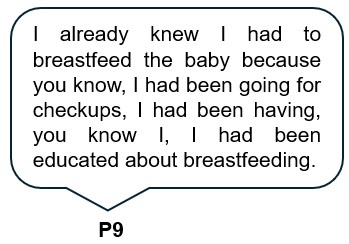 |
| 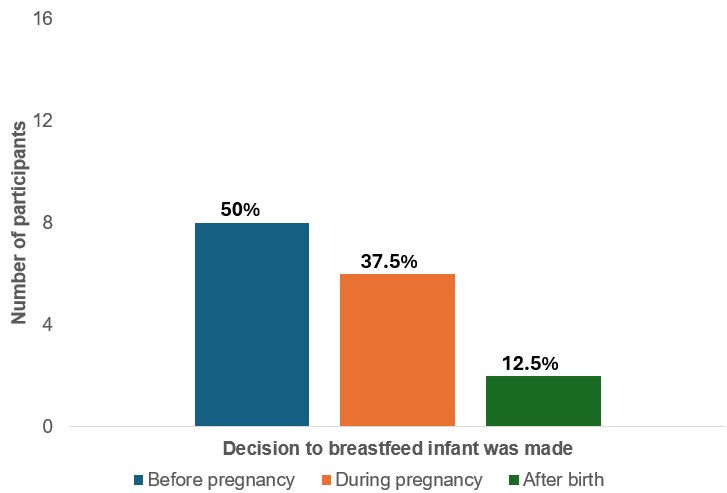 | Subtheme ‘*Decision to breastfeed*’  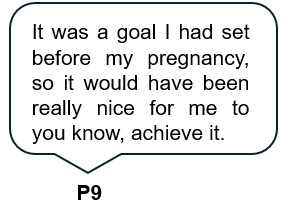 |

In the first quantitative column chart, results were presented for only the 14 participants who answered *yes* to the question “Were you ever depressed at any time after the birth of your child?”
